# Pericoronary adipose tissue radiomics from coronary CT angiography identifies vulnerable plaques characteristics in intravascular OCT

**DOI:** 10.1101/2023.01.09.23284346

**Authors:** Justin N. Kim, Lia Gomez-Perez, Vladislav N. Zimin, Mohamed H. E. Makhlouf, Sadeer Al-Kindi, David L. Wilson, Juhwan Lee

## Abstract

Pericoronary adipose tissue (PCAT) features on CT have been shown to reflect local inflammation, and signals increased cardiovascular risk. Our goal was to determine if PCAT radiomics extracted from coronary CT angiography (CCTA) images are associated with intravascular optical coherence tomography (IVOCT)-identified vulnerable plaque characteristics (e.g., microchannels [MC] and thin-cap fibroatheroma [TCFA]). CCTA and IVOCT images of 30 lesions from 25 patients were registered. Vessels with vulnerable plaques were identified from the registered IVOCT images. PCAT radiomics features were extracted from CCTA images for the lesion region of interest (PCAT-LOI) and the entire vessel (PCAT-Vessel). We extracted 1356 radiomics features, including intensity (first-order), shape, and texture features. Features were reduced using standard approaches (e.g., high feature correlation). Using stratified three-fold cross-validation with 1000 repeats, we determined the ability of PCAT radiomics features from CCTA to predict IVOCT vulnerable plaque characteristics. In identification of TCFA lesions, PCAT-LOI and PCAT-Vessel radiomics models performed comparably (AUC±standard deviation 0.78±0.13, 0.77±0.14). For identification of MC lesions, PCAT-Vessel radiomics model (0.89±0.09) was moderately better associated than that of PCAT-LOI model (0.83±0.12). Both PCAT-LOI and PCAT-Vessel radiomics models also similarly identified coronary vessels thought to be highly vulnerable (i.e., both TCFA and MC) (0.88±0.10, 0.91±0.09). Favorable radiomics features tended to be those describing texture and size of PCAT. PCAT radiomics can identify coronary vessels with TCFA or MC, consistent with IVOCT. CCTA radiomics may improve risk stratification by noninvasively detecting vulnerable plaque characteristics that are only visible with IVOCT.

## Introduction

Atherosclerotic plaque rupture is responsible for most cases of acute coronary syndrome, and thin-cap fibroatheroma (TCFA) is a prevalent precursor of plaque rupture [1], [2]. Microchannels (MC) in plaques are also strong indicators of plaque vulnerability and intraplaque hemorrhage [3], [4]. Early identification of vulnerable plaque characteristics like TCFA and MC is important for risk assessment and treatment planning to improve outcomes. Intravascular optical coherence tomography (IVOCT) is the only imaging modality with sufficient axial resolution (12-18 *µm*) to identify TCFA and MC.

Coronary computed tomography angiography (CCTA) has become a first-line assessment tool for coronary artery disease [5]–[8]. CCTA can assess the plaque burden in the entire coronary tree and identify patients with high-risk plaque characteristics, such as low-attenuation plaque, napkin ring sign, positive remodeling, and spotty plaque calcification, which are associated with future acute coronary syndrome [9], [10]. More recently, the utility of CCTA has been expanded to include assessment of pericoronary adipose tissue (PCAT). PCAT in CCTA images has been further analyzed using various radiomic features, such as intensity, and texture, based on spatial interrelationships of signal intensities [11]. Previous studies have demonstrated that PCAT radiomics can identify myocardial ischemia and predict major adverse cardiovascular events [12]–[14].

In this study, we sought to examine whether PCAT radiomics features are associated with plaque vulnerability assessed by IVOCT. This concurrent approach differs from conventional prediction of far future adverse events, which employs a long history of intervening actions prior to the adverse event. This study is built on our previous studies of IVOCT image analysis [15–17] using the plaque characterization software OCTOPUS [18–22]. Here, we used OCTOPUS [22] to perform semi-automated registration of IVOCT and CCTA images and analyze features both at the lesion and whole vessel levels. We then used a machine learning approach to determine whether PCAT radiomics features from CCTA predict microscopic characteristics of plaque vulnerability (TCFA and MC) seen with IVOCT.

## Methods

We examined the association between CCTA PCAT radiomics and plaque vulnerability as assessed by IVOCT. This concurrent approach differs from conventional prediction of far future adverse events, which employs a long history of intervening actions prior to the adverse event. This study is built on our previous studies of IVOCT image analysis [15]–[17] using the plaque characterization software OCTOPUS [18]–[24]. Here, we used OCTOPUS [22] to perform semi-automated registration of IVOCT and CCTA images and analyze features both at the lesion and whole vessel levels. We then used a machine learning approach to determine whether PCAT radiomics features from CCTA predict microscopic characteristics of plaque vulnerability (TCFA and MC) seen with IVOCT.

### Study Population

This study retrospectively identified patients from the University Hospitals (Cleveland, OH, USA) who underwent both CCTA and IVOCT procedures based on medical indications. Exclusion criteria were as follows: 1) history of myocardial infarction, 2) previous coronary stent implantation, and 3) poor quality of CCTA or IVOCT images. Thirty paired coronary vessel lesions on CCTA and IVOCT were analyzed. This retrospective study was approved by the Institutional Review Board of University Hospitals (Cleveland, OH, USA), conducted following the principles of the Declaration of Helsinki, and the written consent was waived.

### IVOCT imaging

IVOCT images were obtained from a C7XR frequency-domain IVOCT Imaging System (Abbott Vascular, Santa Clara, CA, USA) after an injection of nitroglycerin (100–200 *g*). IVOCT was performed with Dragonfly OPTIS 2.7 F 135-*cm*. Blood clearance was achieved by non-diluted iodine contrast using ISOVUE-370 (iopamidol injection, 370 *mg iodine/mL*; Bracco Diagnostics Inc., Princeton, NJ, USA). The optical probe employed automated pullback at a rate of 36 *mm/s* using survey mode (375 *frames*, 75 *mm*), a frame rate of 180 *frames/s*, and axial resolution of 20 *μm*. IVOCT images were deidentified and analyzed at the Cardiovascular Phenomics Core at University Hospitals. Cardiologist experts with more than 9 years of experience assessed the quality of each pullback for inclusion in the analysis.

### IVOCT processing to extract vulnerability characteristics

Using dedicated OCTOPUS software developed in our previous studies [15]–[24], each coronary vessel was binary-labeled in the presence of TCFA and MC. TCFA was defined as a plaque with the thinnest fibrous cap <65 *µm* and a TCFA angle >90°, and MC was defined as a non-signal tubuloluminal structure without a connection to the vessel lumen, recognized on three or more consecutive cross-sectional IVOCT images [4].

### CCTA acquisition

All CCTA images were acquired on a Brilliance ICT 256 scanner (Philips Healthcare, Cleveland, OH, USA) in accordance with institutional clinical protocols. BMI-specific scanner settings were used (BMI ≤30 *kg/m*^*2*^: 100 *kV/300 mAs*; BMI >30 *kg/m*^*2*^: 120 *kV*/450 *mAs* for prospective gating; and BMI ≤30 *kg/m*^*2*^: 100 *kV*/800 *mAs*, BMI >30 *kg/m*^*2*^: 120 *kV*/800 *mAs*, for retrospective gating), with craniocaudal scan direction. In cases with irregular rhythm, we used retrospective gating without tube modulation. Iodinated contrast dye (ISOVUE 370) 80 mL was injected via 18-gauge angiocath 20 or 22 Diffusics needle (Nexiva™, BD, NJ, USA), preferably, at 5–6 *mL/s* followed by a 70 *mL* saline flush, divided into a 20 *mL* test flush at the rate of 6 *mL*/sec prior to the scan and 50 *mL* bolus-chase after contrast injection. The bolus was tracked in the ascending aorta at the level of the carina with a threshold of 100 HU for optimal imaging. Hyperemia was achieved by sublingual nitroglycerin 5 minutes prior to scan, and a targeted heart rate of 60 or less was achieved using metoprolol 100 *mg* PO and intravenous metoprolol in 5 *mg* increments up to 20 *mg* if systolic pressure was >100 *mmHg*.

Coronary vessels with lesions were multiplanar reformatted on a dedicated workstation (Aquarius Workstation version 4.4.11-13; TeraRecon, Foster City, CA, USA). Then, from the straightened vessel, axial slices of coronary vessels were obtained and stored as DICOM files and sent via secure connection for further PCAT segmentation and analysis.

### PCAT segmentation

PCAT segmentation of each vessel was performed as follows. Vessel walls were segmented automatically using a dedicated workstation. Expert cardiology readers reviewed the results and corrections were made if required. PCAT in CCTA was defined as all voxels located within the radial distance from the outer coronary wall equal to the diameter of the vessel, with CT attenuation values between -190 and -30 HU [13], [25]–[27].

PCAT segmentation was performed semi-automatically using Python software. The software imported the segmented vessel walls, selected the PCAT candidate region per the above definition of PCAT, and thresholded (−190 to -30 HU) to extract the PCAT mask. The PCAT was segmented for each coronary vessel and exported as binary masks for radiomic feature extraction. The average PCAT segmentation time for each vessel was approximately 50 seconds.

### Radiomic analysis

The extracted radiomic features were grouped into three categories—shape, intensity, and texture—defined as follows. Shape-based features describe the shape and geometric properties calculated independently of the gray-level intensity distribution. Intensity (first-order) features depend on the distribution of HU values (e.g., mean, standard deviation, entropy, skewness, kurtosis, etc.) without considering the spatial distribution. Texture features calculate the statistical interrelationship between neighboring voxels [28]. For example, gray-level co-occurrence matrices (GLCM) describe the frequency of co-occurrences of an HU pixel value pair; gray-level dependence matrix (GLDM) quantifies a number of connected voxels within a distance that is dependent on the center voxel [29]; gray-level run length matrix (GLRLM) quantifies the frequency of consecutive occurrences of the same voxel value [30]; gray-level size zone matrix (GLSZM) quantifies homogeneity and variation characteristics, by measuring the number of voxels with the same value [28], [30]; and neighboring gray-tone difference matrix (NGTDM) describes the center and neighboring pixels [30]. For texture feature extraction, voxels were discretized into 8, 16, and 32 bins with identical HU ranges, and all texture-based features were calculated using these three specified bin sizes. Of 1356 radiomics feature extracted, the number of shape, intensity, and texture features were 252, 204, and 900, respectively.

After radiomics feature extraction, to reproduce more features based on the distribution of the extracted radiomics feature values, the minimum, maximum, mean, and standard deviation were calculated. The average time to extract radiomics features and remove highly correlated features for each vessel was approximately 9 minutes. The radiomics features were calculated using the open-source PyRadiomics package in Python [36], which adheres to Image Biomarker Standardization Initiative (IBSI) guidelines [37].

### PCAT radiomics feature extraction

To enable spatial correlative analysis of data, we manually registered IVOCT pullbacks to corresponding vessels in CCTA images by identifying landmark characteristics of the coronary artery such as bifurcations or large calcifications. Two ranges of CCTA frames were selected for PCAT radiomic feature extraction: 1) the CCTA-IVOCT registered plaque lesion of interest, *PCAT*-*LOI*; and 2) the entire set of CCTA frames of the coronary artery, *PCAT-Vessel*. The process to obtain the starting and end point of PCAT-Vessel is as follows. The centerline of artery was automatically identified using Aquarius Workstation (TeraRecon Inc., Foster City, CA, USA) and an expert cardiologist reviewed it. For LAD and LCX, PCAT-Vessel started from the ostium, and for RCA, PCAT-Vessel excluded first 10 *mm* from the ostium. The end point was the end of coronary arteries determined by an expert cardiologist with guidance from the extracted centerline. Segmented PCAT labels from CCTA with their HUs were used for radiomic feature extraction. Each radiomic feature was extracted from respectively selected CCTA frame ranges.

### Association of PCAT radiomics with IVOCT vulnerable characteristics

To determine the associations between CCTA-derived PCAT radiomics and IVOCT vulnerable plaque characteristics, we analyzed the data as follows. Three binary classes were used to represent the vessels with TCFA, MC, and both TCFA and MC (referred to as IVOCT-TCFA, IVOCT-MC, and IVOCT-TCFA-MC, respectively). From the initially extracted 1356 radiomic features from both PCAT-LOI and PCAT-Vessel, highly correlated features (|r| >0.95) were removed. The remaining 293 and 341 radiomic features for PCAT-LOI and PCAT-Vessel, respectively, were used for subsequent analyses.

Univariate analysis was performed to assess diagnostic performance of individual features. For each binary class, each PCAT radiomics feature was fitted to univariate logistic regression using stratified three-fold cross-validation with 1000 repeats. An average receiver operating characteristic (ROC) curve was computed, and the area under the ROC curve (AUC) was used to evaluate the diagnostic performance of each radiomic feature in PCAT-LOI and PCAT-Vessel. The mean AUC of each feature in identification of vessels with TCFA and MC were plotted using a Manhattan plot.

A multivariate analysis was performed to evaluate the overall diagnostic performance of the radiomics model based on feature selection. From the univariate logistic regression, 15 features with the highest mean AUC in each of seven classes of features (shape, first-order, and five subclasses of texture features) were selected. Of the selected 105 features, the minimum redundancy maximum relevance (mRmR) algorithm was used to retain 50 features that had the highest correlation to the output class and the least mutual information among the features, based on the criteria of mutual information difference [33]. The mRmR algorithm further reduced multicollinearity in features that are highly predictive in univariate logistic regression, thereby mitigating overfitting. The final radiomics model was refined to retain 10 features at most, and the optimal set of features was chosen recursively by removing the features of least importance evaluated by the coefficient. The feature selection process was repeated to obtain the PCAT-LOI and PCAT-Vessel radiomics models for the prediction of IVOCT-TCFA, IVOCT-MC, and IVOCT-TCFA-MC. A list of features included the radiomics model is shown in Supplementary Table 1, with the features used in the final multivariate models indicated. The diagnostic performances of models were evaluated with 95% confidence interval of AUC, sensitivity, specificity, positive prediction value, and negative prediction value. The evaluation metrics is shown in Supplementary Table 2. The diagnostic performance of the entire dataset was not generalizable given the limited vessel dataset size. Therefore, stratified three-fold cross-validation with 1000 repeats was implemented to reduce overfitting bias and provide a robust estimate of the expected performance of the models in a new dataset. The mean AUC and standard deviation (SD) of each model were computed to evaluate diagnostic performance. All statistical analyses were done in Python using the Scikit-learn package [34].

## Results

### Patient and lesion characteristics

Table 1 summarizes the baseline characteristics of the study population. Overall, 30 lesions from 25 patients were analyzed. The mean patient age was 63 ± 11 years, and six (26.1%) patients had prior CABG. Most patients were male (75%), had diabetes mellitus (87%) and dyslipidemia (96%). Most lesions were in the left anterior descending artery (LAD; 76.7%). The median (interquartile range [IQR]) time between CCTA and IVOCT procedures was 11 (3–24) days. Of the 30 lesions, IVOCT-TCFA was present in 14, IVOCT-MC in 12, and IVOCT-TCFA-MC in six lesions.

Figures 1A and 1B respectively show the rendering of IVOCT-identified fibrous cap thickness and microchannels along a coronary artery, using OCTOPUS. Fig. 2A shows the outcome of registration of CCTA and IVOCT coronary vessel images by OCTOPUS. The calcified plaque, shown in white, is well-aligned between the imaging modalities. Fig 2B. shows the IVOCT axial slice at non-calcified lesion, and Fig. 2C shows the registered CCTA axial slice at non-calcified lesion, overlayed with PCAT segmentation of the defined HU range.

**Table 1.** Clinical Variables of the Study Population. Values are mean ± standard deviation (SD) for continuous variables, and n (%) for categorical variables. FHx of CAD, family history of coronary artery disease; CABG, coronary artery bypass graft; eGFR, estimated glomerular filtration rate; WBC, white blood cell; LAD, left anterior descending artery; LCx, left circumflex artery, RCA, right coronary artery. *Two missing records.

| Clinical Characteristics | n = 25 |
| --- | --- |
| Age, years | 63 ± 11 |
| Male | 19 (76%) |
| Body Mass Index, kg/m<sup>2</sup> | 28.5 ± 4.7 |
| Cardiovascular Risk Factors |  |
| Hypertension | 11 (44%) |
| Diabetes | 20 (87%*) |
| Chronic Kidney Disease | 11 (44%) |
| Family History of CAD | 14 (56%) |
| Prior CABG | 11 (44%) |
| Current Smoker | 3 (12%) |
| Dyslipidemia | 24 (96%) |
| Blood Parameters |  |
| Creatinine eGFR | 1.5 ± 1.7 |
| WBC count, $\times 10^9/l$ | 15.1 ± 35.4 |
| Hemoglobin, g/dL | 12.9 ± 1.9 |
| Hematocrit, % | 39.4 ± 5.0 |
| Platelet count, $\times 10^9/l$ | 285.5 ± 85.3 |
| HDL-c, mg/dL | 42.1 ± 7.6 |
| LDL-c, mg/dL | 127.0 ± 44.9 |
| Triglycerides, mg/dL | 175.3 ± 88.3 |
| Total-c, mg/dL | 200.3 ± 50.8 |
| Lesion Characteristics | n = 30 |
| Lesion Location |  |
| LAD | 23 (76.7%) |
| LCx | 4 (13.3%) |
| RCA | 3 (10%) |

**Figure 1.**
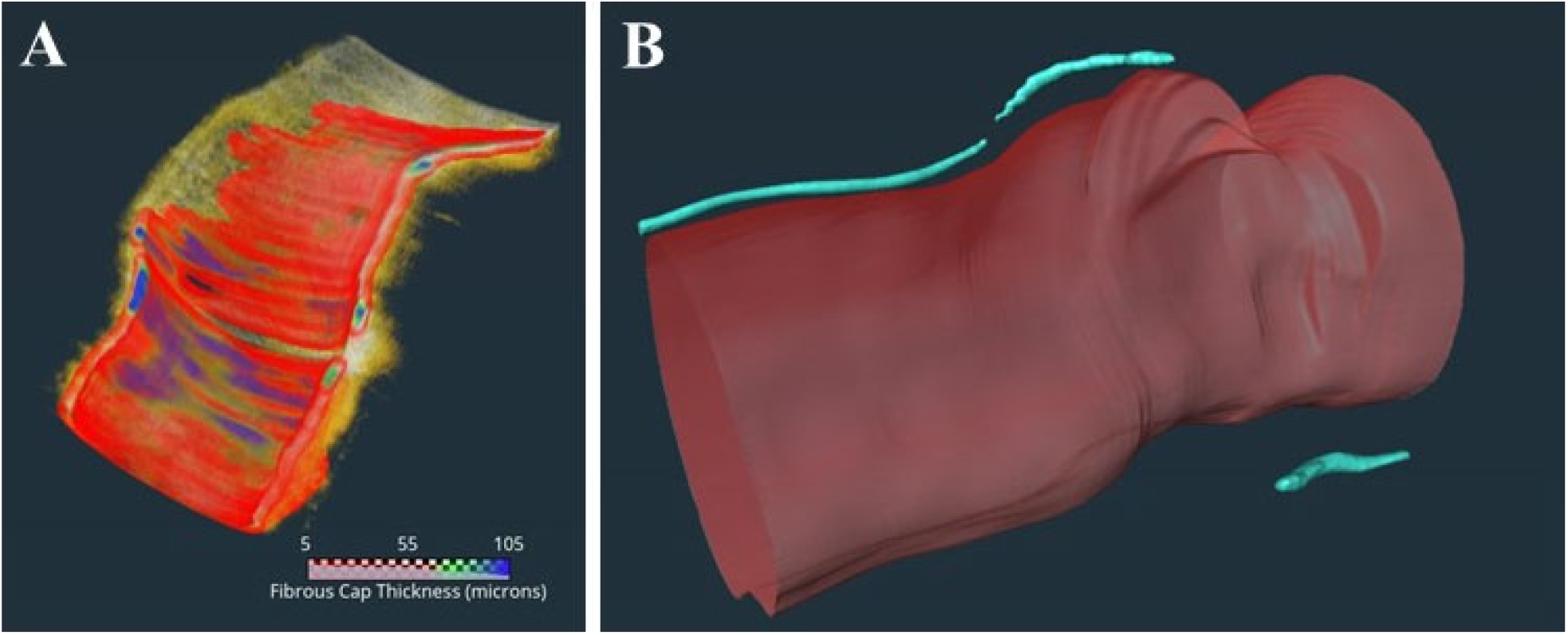
3D visualization of IVOCT coronary artery segments with (A) TCFA and (B) MC. A heatmap of fibrous cap thickness is overlayed in (A), showing the TCFA region. As described in the text, TCFA was defined as a plaque with fibrous cap <65 µm and TCFA angle >90° for each frame. MC was detected as described in the text. In this instance, there are three microvessel (blue) in this plaque. The microvessel segments spans 7.4 mm in length and the diameter is approximately 10.4 µm. Multiple radiomics features captured the extent of TCFA and microvessel presence in a plaque.

**Figure 2.**
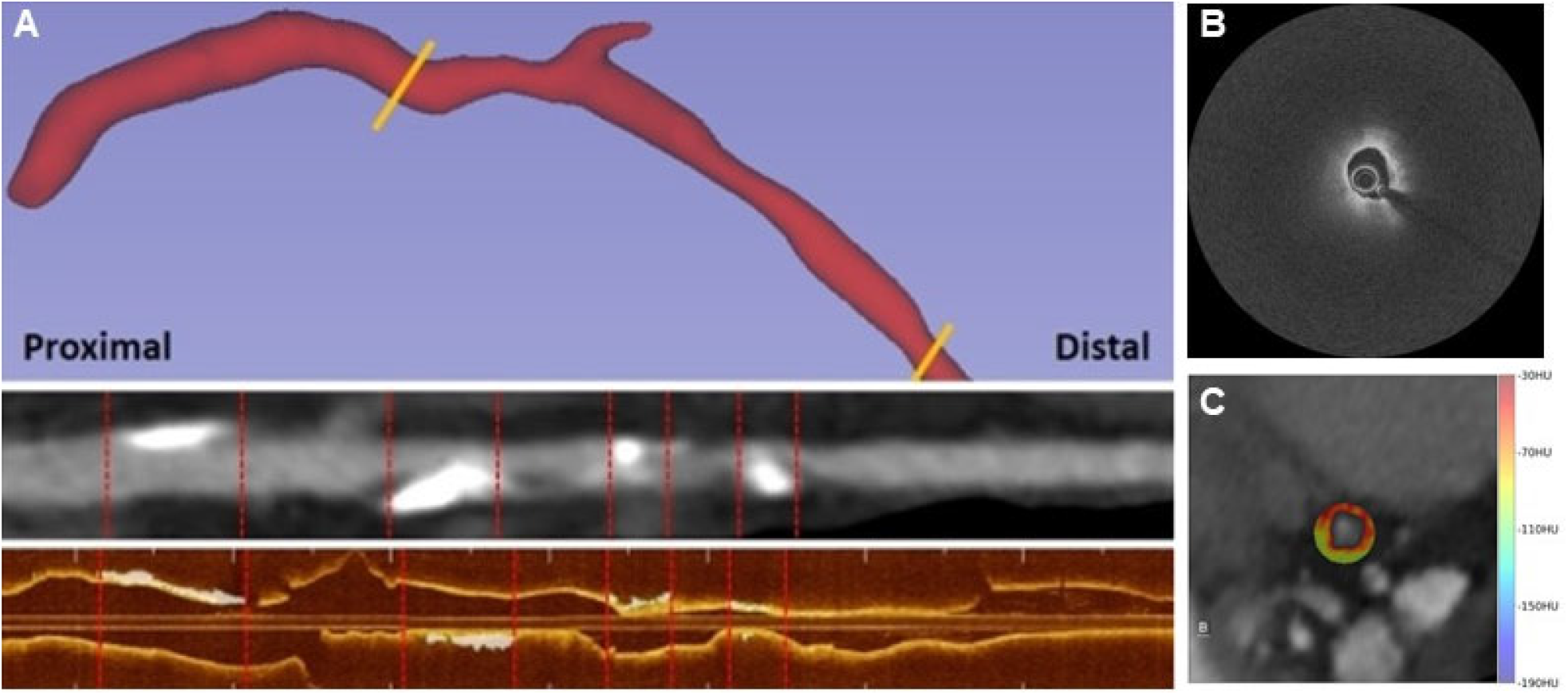
Registration of CCTA and IVOCT. In panel A, we show a 3D CCTA coronary vessel (top), the straightened CCTA (middle), and the IVOCT coronary vessel (bottom). In B, an IVOCT axial frame of non-calcified lesion is shown. In C, a registered CCTA axial frame overlayed with HU colormap segmentation of PCAT is shown. Registration is done by our developed software OCTOPUS. Note that the white calcified plaques in the straightened CCTA view correspond to matching calcifications in IVOCT, demonstrating good registration.

### Univariate analysis of PCAT radiomics to identify IVOCT-TCFA and IVOCT-MC

After removing highly correlated PCAT radiomics features, we analyzed the association of the remaining features with vulnerable plaque characteristics on IVOCT. Of the 293 PCAT-LOI radiomics features (Fig. 3A), six (2.0%) had mean AUC between 0.70 and 0.79, 50 (17.1%) between 0.60 and 0.69, and 91 (31.1%) between 0.50 and 0.59 to identify coronary vessels with IVOCT-TCFA. Of the 341 PCAT-Vessel radiomics features (Fig. 3B), three (3.0%) had mean AUC between 0.70 and 0.79, 26 (26.3%) between 0.60 and 0.69, and 70 (70.7%) between 0.50 and 0.59 to identify coronary vessels with IVOCT-TCFA. The radiomics feature with the highest AUC was the minimum of dependence variance for PCAT-LOI (AUC=0.77, GLDM, discretized at 16 equally sized bins), and the mean of size zone non-uniformity normalized for PCAT-Vessel (AUC=0.73, GLSZM, discretized at 32 bins).

**Figure 3.**
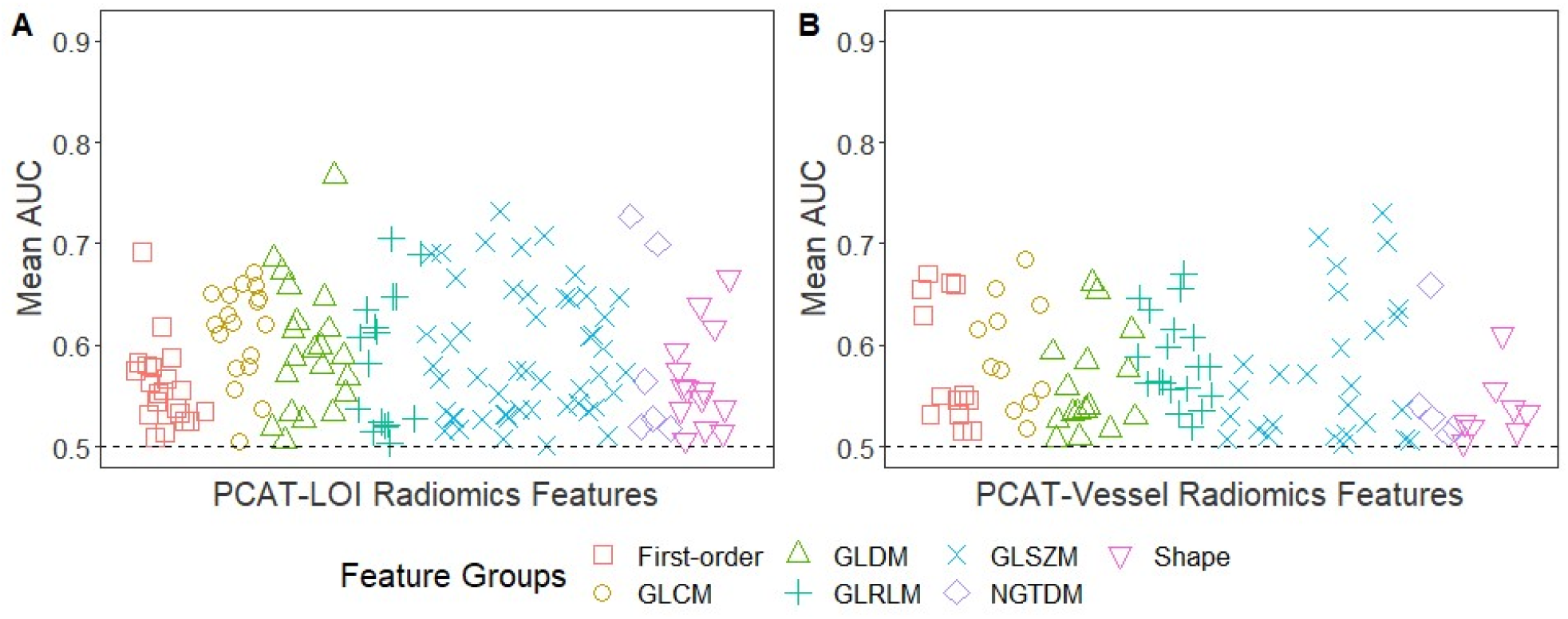
Univariate CCTA feature analysis for predicting IVOCT-TCFA. Manhattan plot of PCAT-LOI (A) and PCAT-Vessel (B) of the AUCs obtained from three-fold cross-validation with 1000 repeats for the identification of coronary vessels with IVOCT-TCFA. The number of radiomics features with AUC >0.5 for identification of coronary vessels with IVOCT-TCFA was 147/293 (50.2%) for PCAT-LOI and 99/341 (29.0%) for PCAT-Vessel. Of the 147 PCAT-LOI radiomics features with AUC >0.5, 14 (9.5%) were shape features, 21 (14.3%) were first-order statistics, and 112 (76.2%) were texture-based features (GLCM: 18, GLDM: 21, GLRLM: 16, GLSZM: 51, NGTDM: 6). Of the 99 PCAT-Vessel radiomics features with AUC >0.5, eight (8.1%) were shape features, 13 (13.1%) were first-order statistics, and 78 (78.8%) were texture-based features (GLCM: 11, GLDM: 16, GLRLM: 19, GLSZM: 27, NGTDM: 5).

The 293 PCAT-LOI radiomics features (Fig. 4A), three (1.0%) had mean AUC between 0.80 and 0.89, 64 (21.8%) between 0.70 and 0.79, 75 (25.6%) between 0.60 and 0.69, and 53 (18.1%) between 0.50 and 0.59 to identify coronary vessels with IVOCT-MC. Of the 341 PCAT-Vessel radiomics features (Fig. 4B), five (1.5%) had mean AUC between 0.80 and 0.89, 37 (10.9%) between 0.70 and 0.79, 82 (24.0%) between 0.60 and 0.69, and 79 (23.2%) between 0.50 and 0.59 to identify coronary vessels with IVOCT-MC. The radiomics features with the highest mean AUC were the maximum of small area high gray-level emphasis (GLSZM, discretized at 8 equally sized bins) for both PCAT-LOI (AUC=0.84) and PCAT-Vessel (AUC=0.89), respectively.

**Figure 4.**
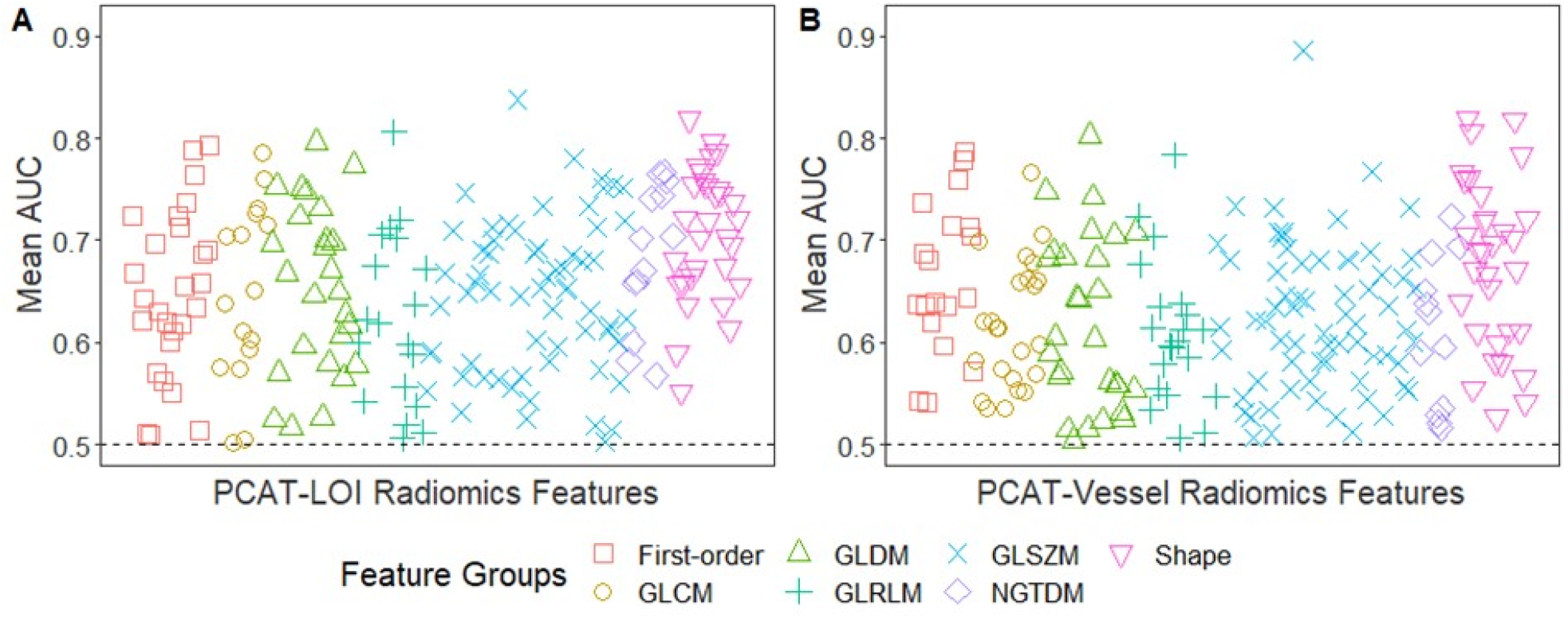
Univariate CCTA radiomics feature analysis for predicting IVOCT-MC. Manhattan plot of PCAT-LOI (A) and PCAT-Vessel (B) of the AUC obtained from three-fold cross-validation with 1000 repeats for the identification of coronary vessels with IVOCT-MC. The number of radiomics features with AUC >0.5 for identification of coronary vessels with IVOCT-MC was 195/293 (66.6%) for PCAT-LOI and 203/341 (59.5%) for PCAT-Vessel. Of 195 radiomics features with AUC >0.5 from the PCAT-LOI, 31 (15.9%) were shape features, 27 (13.9%) were first-order statistics, and 137 (70.3%) were texture-based features (GLCM: 16, GLDM: 26, GLRLM: 19, GLSZM: 63, NGTDM: 13). Of 203 radiomics features from the PCAT-Vessel, 32 (15.8%) were shape features, 19 (9.4%) were first-order statistics, and 152 (74.9%) were texture-based features (GLCM: 24, GLDM: 28, GLRLM: 21, GLSZM: 67, NGTDM: 12).

### Multivariate analysis of PCAT radiomics to identify IVOCT-TCFA and IVOCT-MC

We built radiomic models after a series of feature selection procedures. For the identification of coronary vessels with IVOCT-TCFA, the ROC curves of the PCAT-LOI and PCAT-Vessel radiomics models are plotted in Figure 5A. The PCAT-LOI and PCAT-Vessel models retained three and five radiomic features, respectively. The mean AUC of the PCAT-LOI radiomics model was 0.783 (SD=0.131) and PCAT-Vessel radiomics model was 0.771 (SD=0.135). See Supplementary Table S1 for the final radiomic features used in each model.

**Figure 5.**
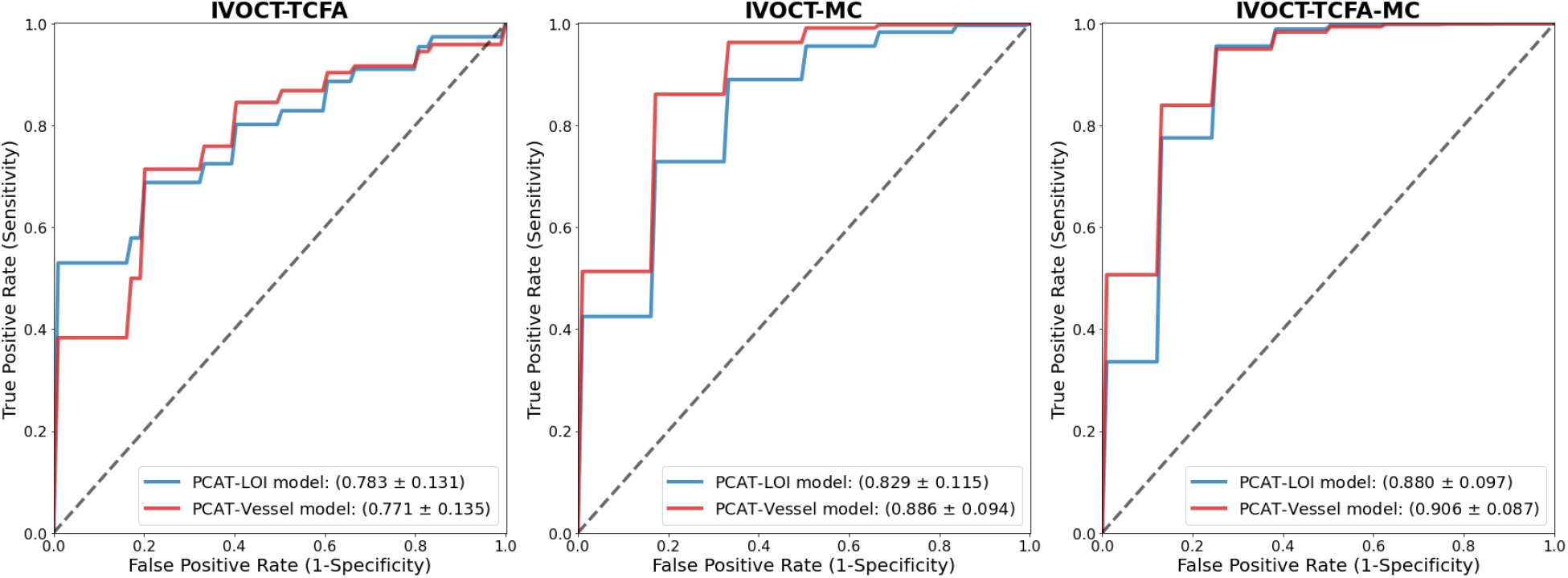
Multivariate analysis of CCTA PCAT radiomics features associated with IVOCT vulnerable plaque characteristics on IVOCT. Diagnostic performance obtained from the three-fold cross-validation with 1000 repeats for the identification of (A) IVOCT-TCFA, (B) IVOCT-MC, and (C) IVOCT-TCFA-MC. ROC curves of PCAT radiomics models for PCAT-LOI and PCAT-Vessel are plotted, and the mean AUC and SD are reported.

For the identification of coronary vessels with IVOCT-MC, the ROC curves of the PCAT-LOI and PCAT-Vessel radiomics models are plotted in Figure 5B. Three radiomics features were retained in the PCAT-LOI and PCAT-Vessel models (Supplementary Table S1). The mean AUC of the PCAT-LOI radiomics model was 0.829 (SD=0.115) and the PCAT-Vessel radiomics model was 0.886 (SD=0.094).

### Radiomic features for identifying vulnerable vessels

Coronary vessels with both TCFA and MC (IVOCT-TCFA-MC) are potentially the most vulnerable plaque and are at high risk of rupture. To evaluate whether PCAT radiomics models could identify these most vulnerable vessels, we repeated the multivariate analysis via feature selection and model building. The PCAT-LOI and PCAT-Vessel models retained three and eight radiomic features, respectively, with no overlapping features in the final models.

Figure 5C shows the ROC curves of the radiomics model in the identification of vulnerable coronary vessels, IVOCT-TCFA-MC. The mean AUC of the PCAT-LOI radiomics model was 0.880 (SD=0.097) and PCAT-Vessel radiomics model was 0.906 (SD=0.087). See Supplementary Table S1 for the final radiomic features used in each model and Supplementary Table S2 for performance evaluation metrics.

## Discussion

Although the limited resolution of CCTA makes it impossible to directly identify plaque microstructures such as TCFA and MC, we were able to identify PCAT features on CCTA images that are associated with microscopic findings of plaque vulnerability in IVOCT. To the best of our knowledge, this is the first study to identify coronary vessels with TCFA and MC using PCAT radiomics.

Non-invasive identification of TCFA and MC may improve risk assessment. TCFA, a precursor of plaque rupture, is the leading cause of acute coronary syndrome [35], and IVOCT-identified TCFA is strongly associated with plaque vulnerability [36], [37]. Previous studies [38]–[40] attempted to differentiate TCFA from non-TCFA lesions on IVOCT using CT attenuation and/or CT-derived high-risk plaque characteristics, but produced conflicting findings. MCs are newly formed microvessels branched from the vasa vasorum, which may promote lipid influx and macrophage infiltration into the coronary plaques [41]. MCs are microstructures in the plaque on IVOCT that is predictive of plaque vulnerability. An increased frequency of MC on IVOCT correlates with a larger frequency of TCFA and positive remodeling [4]. Non-invasive identification of MC in coronary vessels may aid in directing preventative measures to vulnerable plaques.

Our univariate analysis identified several PCAT radiomics features that are predictive of vulnerable plaque characteristics on IVOCT. Both PCAT-LOI and PCAT-Vessel had a similarly large percentage of radiomic features with AUC >0.5 for identifying IVOCT-MC (66.6% and 59.5%, respectively), suggesting that CCTA markers of MC are prevalent both near the lesion and along the entire vessel length. A greater percentage of PCAT-LOI features than PCAT-Vessel features had an AUC >0.5 for identifying IVOCT-TFCA (50.2% vs. 29.0%). This may suggest that PCAT affects the nearby TCFA and corroborates the outside-in theory of atherosclerosis, which proposes that inflamed adipocytes in PCAT contribute to the development of atherosclerosis via the production of adipocytokines. Inflamed adipocytes in PCAT may develop or affect nearby TCFA, and the inflamed PCAT may be captured by a larger percentage of radiomics features. Recently, Chen et al. reported that coronary plaque radiomics, especially wavelets, from CCTA images outperformed conventional high-risk plaque features for identifying IVOCT-derived TCFA [42]. Their plaque radiomics model only comprises of wavelet-filtered features to identify IVOCT-TCFA, but our study demonstrates that PCAT radiomics and its shape identified both IVOCT-TCFA and IVOCT-MC.

In our study, all PCAT radiomics models successfully identified IVOCT-TCFA, IVOCT-MC, and IVOCT-TCFA-MC on CCTA images. Quantitative radiomics analysis of CCTA images may enhance the identification of other microstructures in coronary plaques such as cholesterol crystals and macrophage infiltration. Integration of CCTA radiomics imaging biomarkers can further improve cardiac risk assessment and treatment planning.

Our study has some limitations. This was a retrospective study of a high-risk patient population indicated for IVOCT imaging. In addition, a relatively small number of datasets were obtained from a single center, which may limit generalizability. To account for the limited sample size, AUCs were calculated using stratified three-fold cross-validation with 1000 repeats to enable a determination of robustness across the dataset. All vessel types were combined, with most lesions in the LAD. With larger datasets, each artery could be independently assessed. Further investigations with a larger datasets and multicenter cohorts are needed for further validate our models.

## Conclusion

Our results indicate that non-invasive CCTA-derived PCAT radiomics can identify vessels with vulnerable plaque characteristics (thin-cap fibroatheroma and microchannels) as identified on IVOCT.

## Data Availability

All data produced in the present study are available upon reasonable request to the authors.

## Acknowledgments

This work was supported by the National Science Foundation Graduate Research Fellowship under Grant No 1937968. This project was also supported by the National Heart, Lung, and Blood Institute through grants NIH R21 HL108263, NIH R01 HL114406, and NIH R01 HL143484. This research was conducted in space renovated using funds from an NIH construction grant (C06 RR12463) awarded to Case Western Reserve University. The veracity guarantor, Juhwan Lee, affirms to the best of his knowledge that all aspects of this paper are accurate.

## Supplementary Files

**Table S1.**
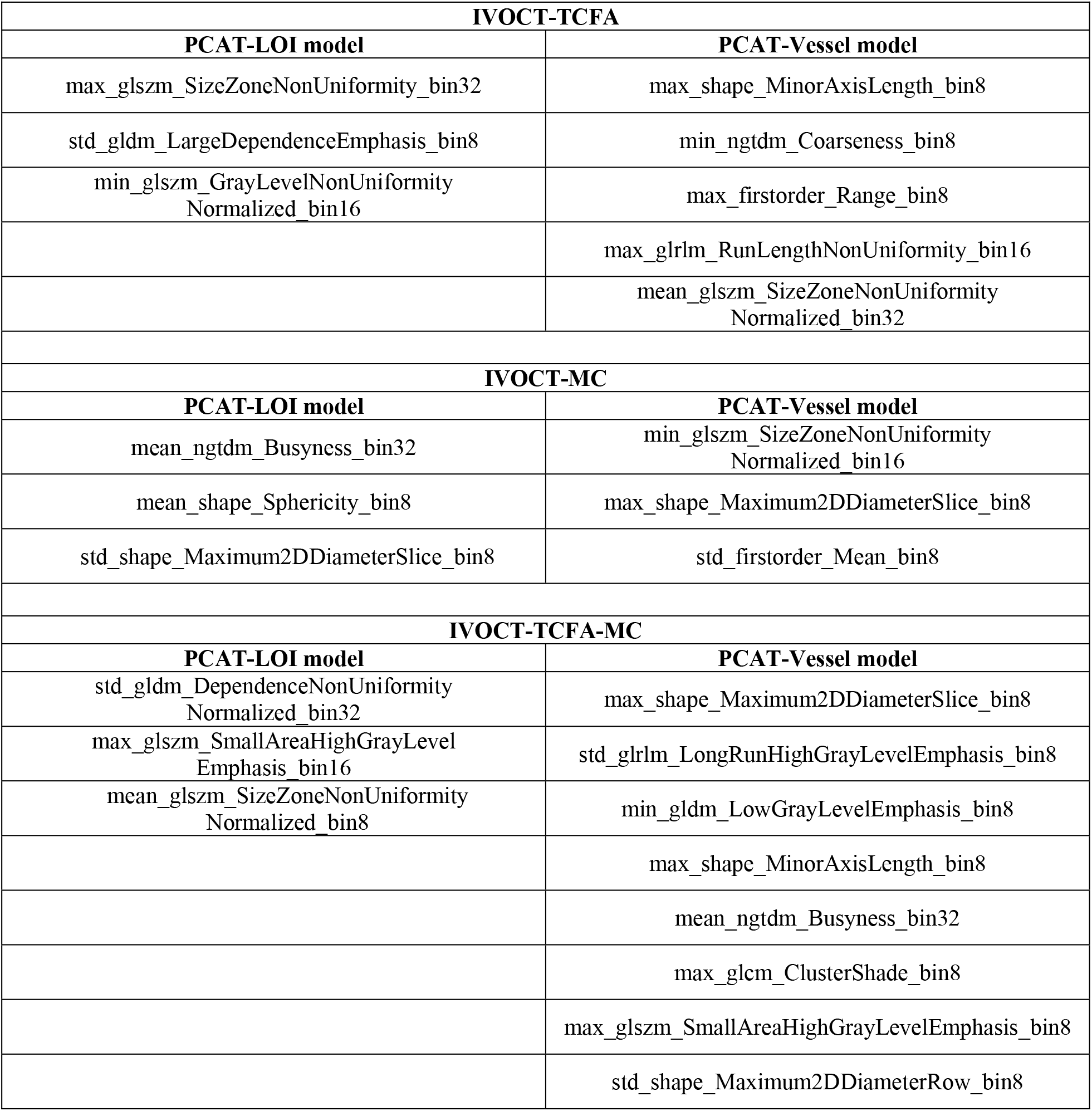
List of features included in the final models of identification of IVOCT-TCFA, IVOCT-MC, and IVOCT-TCFA-MC, respectively. Favorable radiomics features tended to be those describing texture and size of PCAT.

**Table S2.** Evaluation metrices of performance models in identification of IVOCT-TCFA, IVOCT-MC, and IVOCT-TCFA-MC, respectively. TCFA, thin-cap fibroatheroma; MC, microchannel; CI, confidence interval; PPV, positive prediction value; NPV, negative prediction value

| Model | AUC (95% CI) | Sensitivity (95% CI) | Specificity (95% CI) | PPV (95% CI) | NPV (95% CI) |
| --- | --- | --- | --- | --- | --- |
| **IVOCT-TCFA** (Positive/Negative case = 14/16) |  |  |  |  |  |
| **PCAT-LOI radiomics** | **0.783**<br>(0.781-0.787) | 0.656<br>(0.648-0.664) | **0.720**<br>(0.714-0.727) | 0.577<br>(0.569-0.584) | 0.725<br>(0.720-0.731) |
| **PCAT-Vessel model** | 0.771<br>(0.766-0.775) | **0.724**<br>(0.717-0.732) | 0.700<br>(0.693-0.708) | **0.640**<br>(0.632-0.647) | **0.764**<br>(0.758-0.770) |
| **IVOCT-MC** (Positive/Negative case = 12/18) |  |  |  |  |  |
| **PCAT-LOI radiomics** | 0.830<br>(0.826-0.834) | 0.552<br>(0.544-0.560) | 0.821<br>(0.816-0.826) | 0.368<br>(0.362-0.373) | 0.745<br>(0.742-0.749) |
| **PCAT-Vessel model** | **0.886**<br>(0.884-0.891) | **0.636**<br>(0.627-0.644) | **0.859**<br>(0.854-0.863) | **0.424**<br>(0.418-0.429) | **0.796**<br>(0.792-0.800) |
| **IVOCT-TCFA-MC** (Positive/Negative case = 6/24) |  |  |  |  |  |
| **PCAT-LOI radiomics** | 0.880<br>(0.876-0.883) | 0.098<br>(0.09-0.105) | 0.929<br>(0.926-0.932) | 0.024<br>(0.023-0.026) | 0.806<br>(0.805-0.808) |
| **PCAT-Vessel model** | **0.904**<br>(0.901-0.909) | **0.451**<br>(0.439-0.464) | **0.940**<br>(0.937-0.943) | **0.113**<br>(0.110-0.116) | **0.878**<br>(0.875-0.880) |

## Notes

### Competing Interest Statement

The authors have declared no competing interest.

### Author Declarations

This retrospective study was approved by the Institutional Review Board of University Hospitals Cleveland Medical Center (Cleveland, OH, USA), conducted following the principles of the Declaration of Helsinki, and the written consent was waived.

